# Real-world data from the Japanese National Health Insurance System enable fine phenotyping in a 14K-scale population-based study

**DOI:** 10.1101/2025.10.06.25337397

**Authors:** Yayoi Otsuka-Yamasaki, Naoyuki Nishiya, Yoichi Sutoh, Reiko Aoyama, Kozo Tanno, Motoki Nakao, Shohei Komaki, Shiori Minabe, Hideki Ohmomo, Koichi Asahi, Yasushi Ishigaki, Makoto Sasaki, Atsushi Shimizu

**Author notes:** These authors should be considered joint first authors (Yayoi Otsuka-Yamasaki, Naoyuki Nishiya, Yoichi Sutoh). Correspondence: Atsushi Shimizu.

## Abstract

The use of medical databases, known as real-world data (RWD), enables accurate and efficient estimation of disease prevalence in population-based studies, making it a potential game-changer in epidemiology. However, the lack of standardized data formats across hospitals complicates integration across institutions. In Japan, health insurance claims data are standardized under a nationally unified format, providing a reliable source of structured RWD. We evaluated the utility of insurance claims data in epidemiological research. Incorporating both diagnosis and prescription information into case definitions resulted in four- and six-fold increases in the estimated prevalence of Alzheimer’s disease (AD) and Parkinson’s disease (PD), respectively, compared with conventional self-reported definitions. Subsequent genome-wide association studies (GWAS) for AD showed increased model log-likelihood and identified a characteristic *APOE* signal, findings observed only with extended case definitions. The *APOE* effect size was consistent with large case–control studies, while standard errors remained comparable to smaller studies. These results indicate that claims-based phenotyping improves case identification without loss of accuracy and supports scalable approaches for genomic epidemiology and public health surveillance.

**Patient consent statement:** All participants provided written informed consent and the study protocol was approved by the Institutional Review Board of Iwate Medical University (Approval number HG2021-009)

**Permission to reproduce material from other sources:** No materials requiring permission from other sources have been used in this manuscript.

**Clinical trial registration:** This study does not involve interventional components and therefore was not registered as a clinical trial.

## Introduction

The accurate estimation of disease prevalence within a community is crucial for public health. However, challenges remain. Recent advances in medical databases—such as electronic health records and health insurance claims data, collectively referred to as real-world data (RWD)—provide opportunities to enhance the accuracy, efficiency, and frequency of prevalence estimation in population-based studies. These developments also enable the scale-up of surveys and inform public health decision-making (Liu & Panagiotakos, 2022; Swift et al., 2018). Furthermore, the integration of RWD with large-scale genome cohorts increasingly contributes to the generation of epidemiological evidence that informs genomic research (Briggs et al., 2022). Nonetheless, technical challenges remain. Although diagnoses recorded in electronic health records are generally reliable, the lack of standardized data formats across hospitals in many regions limit the integration of data across institutions (Hiramatsu et al., 2021).

Insurance claims data offer a standardized format that facilitates integration across institutions, particularly in countries with universal health coverage, such as Japan. In Japan, national health insurance claims are standardized across diverse healthcare providers and administered by local governments (Shida et al., 2023). The Japanese health insurance system consists of two major schemes: the National Health Insurance (NHI), which covers individuals aged ≤74 years who are not enrolled in employee insurance, and the Late-stage Medical Care System for the Elderly (LsMCSE), which covers individuals aged ≥75 years (Ministry of Health, Labour and Welfare., 2025). Individuals are typically insured through their local municipal governments. Both systems provide structured diagnostic and prescription claims data, consequently enabling the retrieval and integration of health data from multiple regions under appropriate legal and ethical frameworks (Fujinaga & Fukuoka, 2022).

The Tohoku Medical Megabank Community Cohort (TMM CommCohort) is a large-scale population-based cohort study established to monitor public health in areas affected by the 2011 Great East Japan Earthquake (Kuriyama et al., 2016). This cohort obtained health insurance claims data through agreements with participating local governments. In this context, diagnosis and prescription data from claims can supplement medical histories based on self-administered questionnaires (SAQs) and serve as proxies for clinical diagnoses.

In this study, we evaluated the incorporation of diagnosis and prescription data from insurance claims into conventional case–control diagnostic definitions. The prevalence of hypertension, dyslipidemia, diabetes, Parkinson’s disease (PD), and Alzheimer’s disease (AD) is expected to increase in aging societies. Therefore, we aimed to determine whether incorporating diagnosis and prescription data from insurance claims could improve case– control definitions for these common diseases. We further sought to evaluate the accuracy of phenotyping using an extended case definition incorporating claims-based diagnosis and prescription records in the context of genome-wide association studies (GWAS). These findings may facilitate the development of more scalable strategies in genomic epidemiology and public health research. In addition, they highlight the potential value of a feedback cycle in which claims data inform both public health action and evidence-based policy development for insured individuals.

## Materials and Methods

### Population

The design of the TMM CommCohort study was described previously (Kuriyama et al., 2016). In the present study, we included participants from the TMM CommCohort study who held insurance from the NHI or LsMCSE and resided in Iwate Prefecture. Individuals with missing genotypic, phenotypic, and claims data, or other data necessary for analysis, were excluded from the study.

Participants were recruited during specific health checkups or through visits to cohort assessment centers between May 2013 and March 2016. Medical history (e.g., hypertension, dyslipidemia, diabetes, AD, and PD diagnoses) and treatment status for selected conditions (hypertension, dyslipidemia, and diabetes) were collected using SAQs. Systolic and diastolic blood pressure were measured twice at recruitment, and the average was used for analysis. Hemoglobin A1c (HbA1c), high-(HDL) and low-density lipoprotein (LDL) cholesterol, and triglyceride levels were measured in blood samples collected at recruitment.

National health insurance claims data, including medical, pharmacy, and Diagnosis Procedure Combination receipts, were collected annually by electronically collating with participant information from the TMM CommCohort study in Iwate. This process enabled the construction of a claims-based dataset for TMM participants in Iwate. Personally identifiable information and non-research related elements were excluded to ensure strict data minimization and privacy protection. The claims data are structured in a standard format used across Japan’s public health insurance systems. Clinical endpoints were defined by a combination of disease and medication.

This study adhered to the principles of the Declaration of Helsinki, and written informed consent was obtained from all participants. All individual data were anonymized before analysis. The study was approved by the Institutional Review Board of Iwate Medical University (Approval number HG H25-2). Analyses involving personal genotypic or phenotypic information were performed on a standalone computing system—the Tohoku University Tohoku Medical Megabank Organization supercomputer.

### Genotyping and GWAS

Genotyping and genotype imputation were performed as previously described (Minegishi et al., 2019; Sutoh et al., 2025; Yasuda et al., 2019). In the hard-call genotype data, variants that met any of the following criteria were excluded: low call rate (<0.99), low heterozygous allele count (<5), deviation from Hardy–Weinberg equilibrium (*P* < 1 × 10-6), low minor allele frequency (<0.01), or location within regions of strong linkage disequilibrium (Price et al., 2008). Variant filtering was conducted using PLINK v1.9 (version b5.1) (Chang et al., 2015). Linkage disequilibrium pruning was performed with PLINK v2.0 (version a3LM) using the option “--indep-pairwise 1500 150 0.03”.

Principal component analysis was conducted in PLINK v2.0 using the “--pca approx allele-wts” option. One individual from each kinship pair (kinship > 0.0884) was excluded based on the KING method to account for relatedness (Manichaikul et al., 2010). The resulting principal components were then projected onto all individuals, including those excluded based on the kinship threshold.

Case–control GWAS were conducted using SAIGE v0.44.6.5 (Zhou et al., 2018). Individuals with a low call rate (<0.99) or extreme genetic ancestry, defined as values of PC1 or PC2 deviating by ≥4 standard deviations from the mean, were excluded. In addition, imputed variants with low imputation quality (*R^2^* < 0.3) or allele frequency (<0.01) were excluded from the analysis.

### Statistical analysis

Statistical analyses were performed using R versions 4.1.0 and 4.3.2. Meta-analysis was performed with the R package "meta" (version 8.0-2) (Schwarzer et al., 2015).

### Use of artificial intelligence tools

During the preparation of this work, the authors used ChatGPT 4o (OpenAI, version 4o, including Deep Research mode) to assist in reviewing and summarizing previous studies and improve the computer scripts for data analysis. The authors subsequently verified and edited all content generated by this tool and take full responsibility for the integrity and accuracy of the final manuscript.

## Results

### Retrieval of health insurance claims data and phenotyping

A workflow of the study is shown in Figure 1.

**Figure 1.**
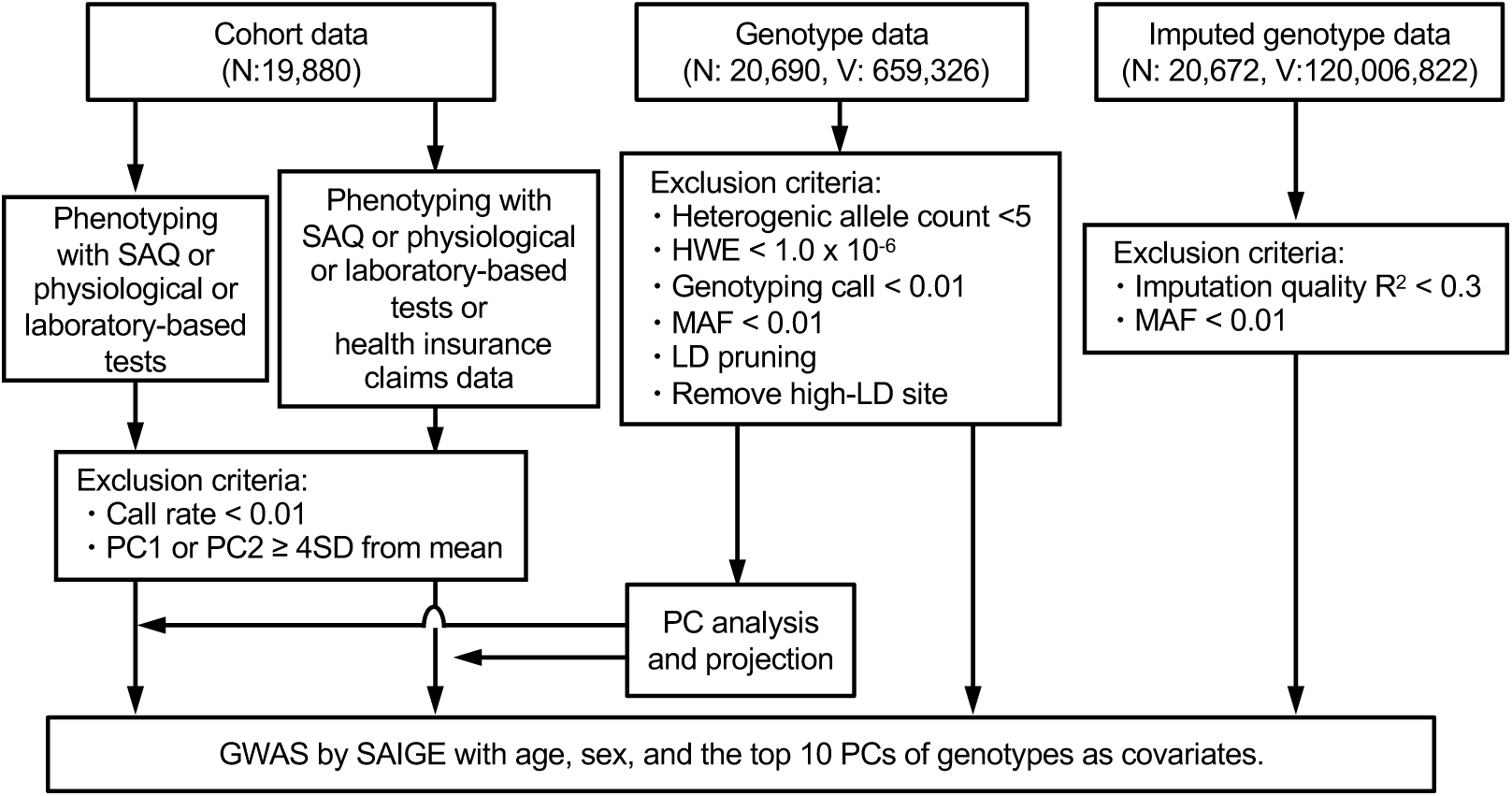
The workflow in this study. The study participants had hard-called and imputed genotype data, as well as cohort information including health insurance claims, demographic characteristics (age and sex), laboratory test results, and responses to pre-specified items in a self-administered questionnaire (SAQ). The real-world data used in this study included disease diagnoses and prescription histories derived from medical, pharmacy, and Diagnosis Procedure Combination claims data. Phenotyping was performed using either conventional definitions based on SAQs and laboratory tests or extended definitions incorporating health insurance claims data. HWE, Hardy-Weinberg equilibrium; LD, linkage disequilibrium; MAF, minor allele frequency; PC, principal component; SD, standard deviation; V, variants.

Health insurance claims data on diagnosis and drug prescriptions were obtained for 19,880 individuals (7,638 men and 12,242 women) (Figure 1). Of these, 14,241 participants met the cohort information and genotype data requirements. Claims data for hypertension, diabetes, and dyslipidemia were retrieved from the NHI system and covered the period between June 2013 and December 2019. The claims data for AD and PD were collected between June 2013 and December 2020 and included data from both the NHI and LsMCSE. Participants were classified into case and control groups using two criteria: (1) the conventional case definition based on SAQs and laboratory tests used in the cohort study, and (2) an extended case definition incorporating both disease names recorded in claims and drug prescription data, in addition to (1) (Table 1).

**Table 1.**
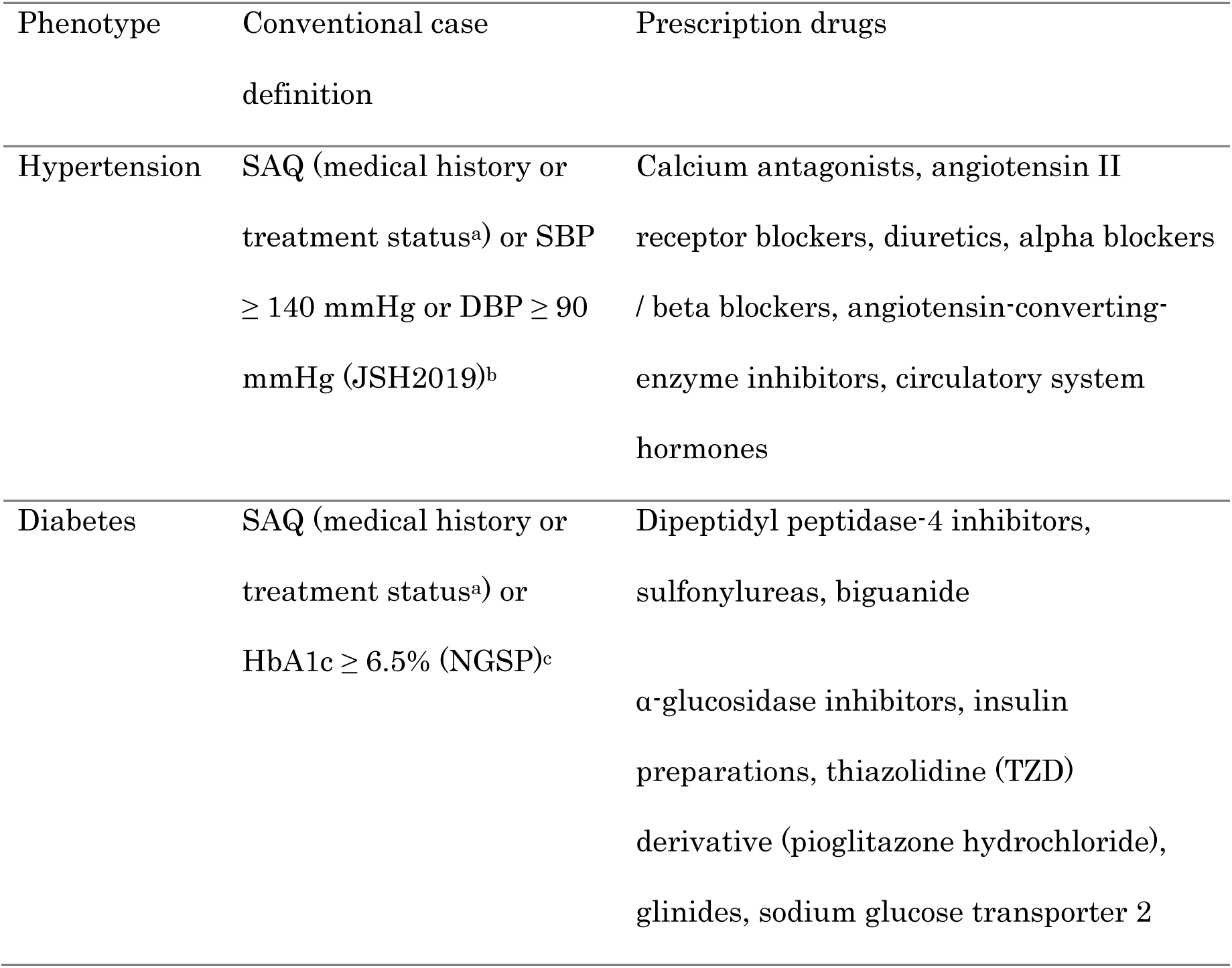
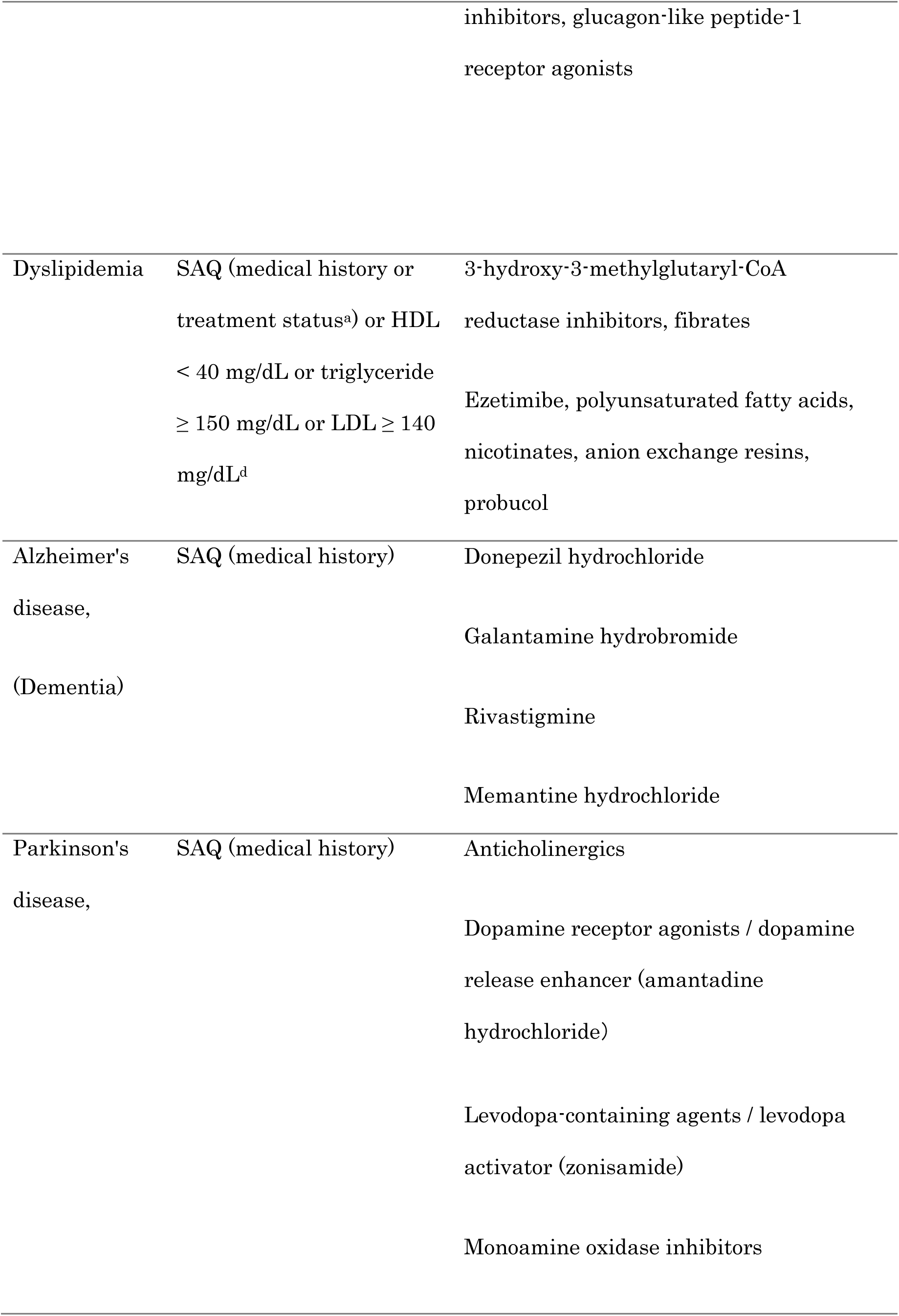
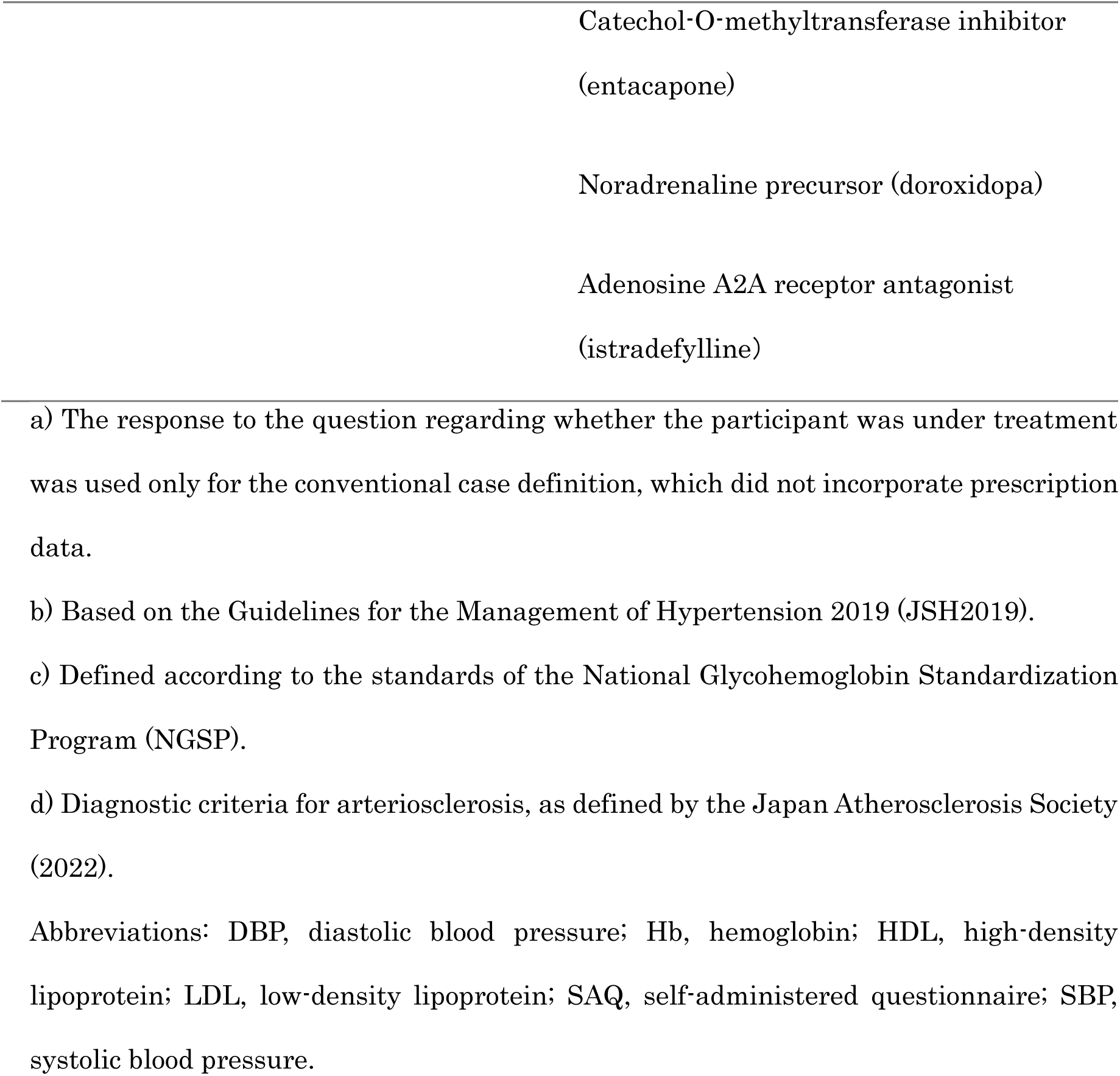
Case definition and prescription drugs for target outcomes.

The incorporation of claims data into phenotyping increased the prevalence of all target outcomes by 1.0–6.3% (Table 2). Common lifestyle-related diseases such as hypertension, diabetes, and dyslipidemia showed modest increases of 4.0%, 1.0%, and 6.3%, respectively. In contrast, the prevalence of AD and PD increased by more than six- and four-fold, respectively.

**Table 2.** Characteristics and prevalence by phenotype.

|  |  |
| --- | --- |
| N | 14,241<sup>a</sup> |
| Female (%) | 61.5 |

Age 61.9 +/- 10.0
|  | Conventional case definition | +claims data |
| --- | --- | --- |
| Hypertension (% (n)) | 46.7 (6,644) | 50.6 (7,211) |
| Diabetes (% (n)) | 11.2 (1,599) | 12.2 (1,740) |
| Dyslipidemia (% (n)) | 53.6 (7,362) | 59.9 (8,531) |
| Alzheimer's disease (% (n)) | 0.2 (22) | 1.3 (183) |
| Parkinson's disease (% (n)) | 0.2 (34) | 0.7 (106) |

a) One participant was missing from the analysis of Alzheimer's disease, Parkinson's disease, and dyslipidemia.

### GWAS

The effects of different phenotyping methods on GWAS results were confirmed (Figure 2A).

**Figure 2.**
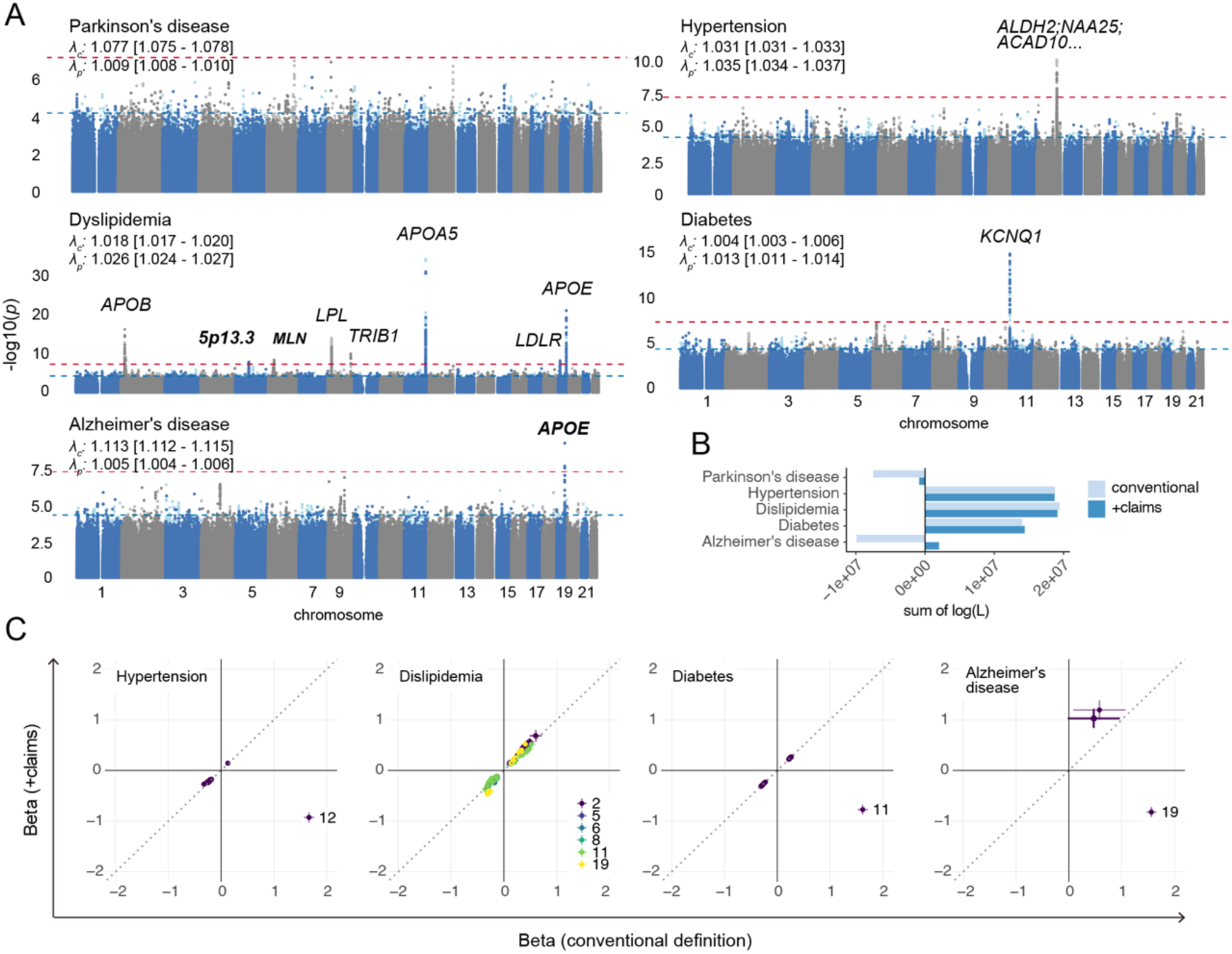
Genome-wide association studies (GWAS) using two case definitions. GWAS were performed using two different case definitions: (1) the conventional method based on SAQs and laboratory tests (light-colored Manhattan plot; most of the plot is hidden), and (2) the extended method incorporating health insurance claims data (dark-colored Manhattan plot) (A). Gene names near genome-wide significant variants are shown in italics; those identified only in the GWAS using definition (2) are shown in bold italics. The red and blue dashed horizontal lines indicate the thresholds for genome-wide significance (*P* = 5 × 10^-8^) and suggestive significance (*P* = 5 × 10^-5^), respectively. *λc/λp* denote genomic inflation factors for the GWAS based on case definitions (1) and (2), respectively; 95% confidence intervals are shown in square brackets. Comparison of the sum of log-likelihoods (log L) from GWAS summary statistics between the two case definitions (B). The number of variants and sample sizes were identical between the two analyses. Effect sizes and directions of significant variants identified in GWAS using the two different case definitions were compared (C). Variants are color-coded by chromosome. Bars and points represent the standard errors and mean of the beta values, respectively. The dotted line represents the diagonal.

Several genome-wide significant signals (*P* < 5 × 10^-8^) were detected for hypertension, dyslipidemia, diabetes, and AD (Figure 2A and Supplementary Tables 1–7). *ALDH2* and *KCNQ1* were identified in the GWAS for hypertension and diabetes. Both genes have been reported for blood pressure and HbA1c, respectively, in previous GWAS (Sakaue et al., 2021). Known lipid-associated loci such as *APOB, LPL, TRIB1, APOA5, LDLR*, and *APOE* were replicated in the GWAS for dyslipidemia, consistent with prior findings on LDL HDL, total cholesterol, and triglyceride levels. In contrast, association signals at 5p13.3 and the motilin gene locus were detected only in the GWAS based on the extended case definition (Figure 2A). These two genes have been detected in the LDL cholesterol or triglyceride GWAS of the TMM CommCohort study, which included 67K participants (Study ID: TGA000007, https://jmorp.megabank.tohoku.ac.jp/gwas-studies/) and Asian populations (Chen et al., 2023; Hsu et al., 2025; Ismail Umlai et al., 2025). The lead variant rs4704208 in the 5p13.3 region identified in this study is an expression quantitative trait locus for the 3-hydroxy-3-methylglutaryl-CoA reductase gene (Figure 2A), as determined according to the GTEx database (https://www.gtexportal.org/home/). This enzyme is the pharmacological target of statins, which are widely prescribed for the treatment of dyslipidemia.

The GWAS for AD using the extended case definitions revealed significant signals in *APOE*, whereas these signals were not detected in the GWAS based on conventional phenotyping (Figure 2A and Supplementary Table 7). The log-likelihood values, which indicate model fit, increased with the use of the extended definition for AD (Figure 2B). The effect sizes and directions of significant variants were consistent across the different phenotyping definitions (Figure 2C). For AD, using the extended case definition yielded smaller standard errors of effect size estimates than the conventional definition, indicating improved estimation precision.

### Comparison with previous studies

The rs429358 C allele, a missense variant in *APOE* that results in a cysteine-to-arginine substitution, is classified as a risk and/or pathogenic allele for AD (ClinVar accession: VCV000017864.36). This variant was replicated in the present GWAS for AD (Figure 2A and Supplementary Table 7). In total, 976 studies reported rs429358-associated hits in the GWAS catalog (Cerezo et al., 2025). Of these, seven that reported associations with AD (Moreno-Grau et al., 2019; Wang et al., 2021; Nazarian et al., 2019; Hong et al., 2020; Akgun et al., 2024; Harper et al., 2022; Li et al., 2021) (Supplementary Table 8), mostly based on clinically diagnosed cases using established guidelines, were comparable to the present study (Figure 3).

**Figure 3.**
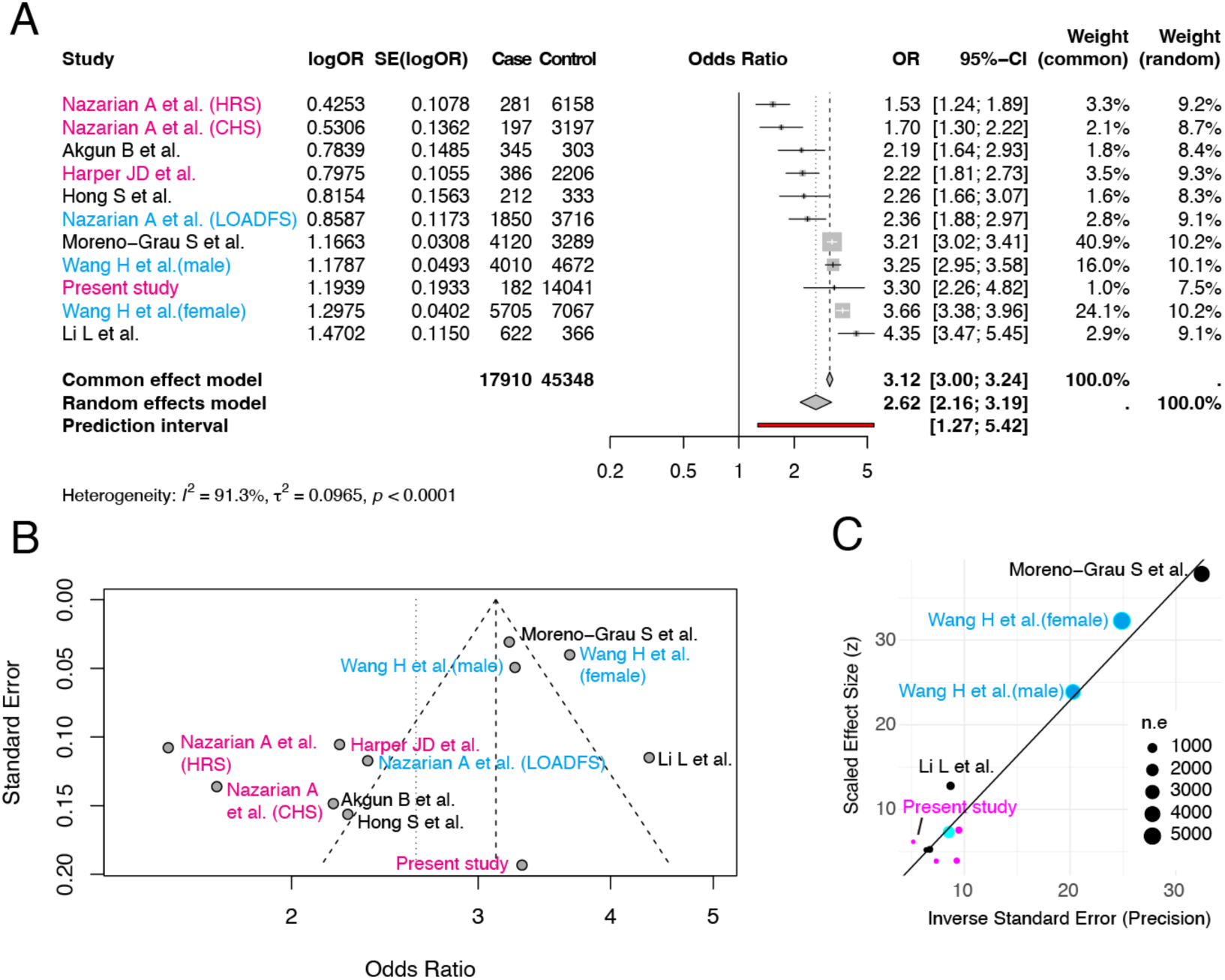
Comparison with previous results. Effect sizes of rs429358 were compared with those reported in previous studies registered in the GWAS Catalog (Cerezo et al., 2025) (A). Studies associated with the trait “Alzheimer’s disease” were selected. The effect size for each cohort (or subgroup) was used when available for analysis. The study design is indicated by color: magenta for population-based studies, black for case–control studies, and cyan for other designs (including meta-analyses and family-based studies). A funnel plot (B) and Egger’s test (C) were used to assess potential publication bias. The linear regression test for funnel plot asymmetry yielded an intercept of -3.49 ± 1.52 (*P* = 0.0484). HRS, University of Michigan Health and Retirement Study; CHS, Cardiovascular Health Study; LOADFS, Late-Onset Alzheimer’s Disease Family Study.

The odds ratio estimated in our study was 3.30 (95% CI: 2.26–4.82). This is consistent with estimates from larger-scale studies (number of cases > 4000; 3.21 (3.02–3.41) reported by Moreno-Grau et al (Moreno-Grau et al., 2019) and Wang et al (Wang et al., 2021) (3.25 (2.94–3.58) and 3.66 (3.37–3.97)) (Figure 3A, B). Moreover, the precision in our study was similar to that in studies with smaller cases (number of cases < 500), such as those by Nazarian et al (Nazarian et al., 2019) and Hong et al (Hong et al., 2020) (Figure 3C). Egger’s test revealed a significant asymmetry (-3.49 ± 1.52; *P* = 0.0484) in the effect size estimates of these GWAS.

## Discussion

According to the NIA-AA research framework (Jack et al., 2018), a biological definition of AD requires costly and time-consuming assessments, such as biomarker testing using positron emission tomography or cerebrospinal fluid analysis. However, increasing case numbers and frequency of follow-up limit the practical application of these approaches. The more than six-fold increase in the estimated prevalence of AD demonstrates the effectiveness of RWD-based phenotyping in community-based cohort studies.

In the present study, the estimated prevalence of AD in the cohort was 1.3%. By 2025, the prevalence of dementia is estimated to be 1.10% (95% CI: 1.07–1.17) in men and 3.47% (95% CI: 3.41–3.52) among women aged 60–74 years in Japan (Kasajima et al., 2022). As women comprised 61.5% of our dataset, the expected overall prevalence of dementia was approximately 2.56%, calculated as 0.615 × 0.0347 + 0.386 × 0.011. Assuming that AD accounts for approximately two-thirds of all dementia cases in Japan (Ikeda et al., 2021; Montgomery et al., 2018), its estimated prevalence would be approximately 1.7%, resulting in a smaller gap with the estimate derived using RWD. However, our dataset included claims data from the LsMCSE, thereby covering individuals aged 75 years and older. Because the prevalence of AD increases substantially with age (Kasajima et al., 2022)— especially among individuals aged ≥75—the inclusion of older adults may increase the overall prevalence. Nevertheless, given that the participants were followed up for 7 years, and most of the cohort remained under 75 years old, the observed prevalence of 1.3% may still represent a reasonable estimate.

Case–control classification based on RWD also affected the GWAS results for AD. The increased log-likelihood values suggest that this updated phenotyping approach may reduce the effect of unmeasured confounders that are not included in the regression models. The replicated detection of the *APOE* variant rs429358, which has been identified as an AD risk allele in several studies, further supports the validity of this approach. The odds ratio estimated in the present study was similar to those reported in large-scale clinical or meta-analytic GWAS with more than 4,000 cases. However, the standard error remained comparable to those from studies with similar case numbers. These findings suggest that the revised case definition improved case identification while maintaining accuracy.

The prevalence of PD also increased when claims data were included. However, no significant improvement was observed in the GWAS results. Although a few studies have reported the prevalence of PD in Japan, regional estimates for East Asia suggested an all-age prevalence of 0.357% (95% CI: 0.287–0.428) in 2021, which is estimated to increase to 0.821% (95% CI: 0.674–0.995) by 2050 (Su et al., 2025). The estimated prevalence of 0.7% in the present cohort, which is predominantly older, is consistent with these projections. A limitation of this study is the potential selection bias due to the characteristics of the insured population. This study included claims data from the NHI system, which covers approximately 30% of the Japanese population, and from the LsMCSE system for those aged 75 years and older. Therefore, it does not represent the entire population (Shigeoka, 2014). Additionally, we could not directly compare the phenotypes estimated from prescription data with clinical diagnoses made in hospitals. This was due to ethical constraints and the substantial human effort required to collect diagnosis data from the hospitals where cohort participants were treated. However, we plan to address this limitation in future research.

In conclusion, this study demonstrates that incorporating RWD into case definitions can improve the efficiency of epidemiological research in population-based studies while maintaining accuracy. These approaches may improve public health surveillance and contribute to the advancement of precision medicine.

## Data Availability

The GWAS summaries have been published in jMorp (https://jmorp.megabank.tohoku.ac.jp/gwas-studies/TGA000016). The datasets of individuals in the TMM CommCohort study analyzed here are not publicly available owing to ethical reasons, such as the protection of personal information and prevention of unintended personal identification. However, they are accessible upon request, subject to approval from the Ethics Committee of Iwate Medical University and the Materials and Information Distribution Review Committee of the TMM Project. Those interested in obtaining the datasets for individuals may contact the corresponding author.

## Data Availability Statement

The GWAS summaries have been published in jMorp (https://jmorp.megabank.tohoku.ac.jp/gwas-studies/TGA000016). The datasets of individuals in the TMM CommCohort study analyzed here are not publicly available for ethical reasons, such as protecting personal information and preventing unintended personal identification. However, they are available from the corresponding author upon request, subject to approval from the Ethics Committee of Iwate Medical University and the Materials and Information Distribution Review Committee of the TMM Project.

## Ethical approval

This study was approved by the Institutional Review Board of Iwate Medical University (Approval number HG2021-009).

## Supplementary information

Supplemental_Tables.xlsx. The Excel file includes Supplementary Tables 1–8.

## Acknowledgements

This research utilized the supercomputer system and dbTMM provided by the Tohoku Medical Megabank Project, supported by AMED under grant numbers JP21tm0424601 and JP21tm0124005.

## Author contributions

Conceptualization: YO-Y, YS, AS; Methodology: YO-Y, YS, AS; Investigation: NN, RA, KT; Formal analysis: YO-Y, NN, YS; Data curation: YO-Y, NN, YS, KT, AS; Writing—original draft: YO-Y, NN, YS; Writing—review & editing: YO-Y, NN, YS, KT, MN, SK, SM, HO, KA, YI, MS, AS.

